# Joint estimation of relaxation and diffusion tissue parameters for prostate cancer grading with relaxation-VERDICT MRI

**DOI:** 10.1101/2021.06.24.21259440

**Authors:** Marco Palombo, Vanya Valindria, Saurabh Singh, Eleni Chiou, Francesco Giganti, Hayley Pye, Hayley C. Whitaker, David Atkinson, Shonit Punwani, Daniel C. Alexander, Eleftheria Panagiotaki

**Affiliations:** Centre for Medical Image Computing, Department of Computer Science, University College London, London (UK); Cardiff University Brain Research Imaging Centre, School of Psychology, Cardiff University, Cardiff (UK); School of Computer Science and Informatics, Cardiff University, Cardiff (UK); Centre for Medical Imaging, University College London, London (UK); Division of Surgery and Interventional Science, University College London, London (UK); Department of Radiology, University College London Hospital NHS Foundation Trust, London (UK); Molecular Diagnostics and Therapeutics Group, Division of Surgery & Interventional Science, University College London, London, (UK)

**Keywords:** biophysical modelling, relaxation-diffusion MRI, deep learning, convolutional neural network, prostate cancer diagnosis, prostate cancer grading

## Abstract

**Purpose:** The Vascular, Extracellular and Restricted Diffusion for Cytometry in Tumours (VERDICT) technique has shown promise discriminating normal from prostate cancer (PCa) tissue and Gleason grade 3+3 from 3+4. However, VERDICT currently doesn’t account for the inherent relaxation properties of the tissue that could add complementary information and potentially enhance its diagnostic power. The aim of this work is to introduce relaxation-VERDICT (rVERDICT) for prostate, a model for the joint estimation of diffusion and relaxation parameters.

**Methods:** 72 men with suspected PCa underwent multiparametric MRI (mp-MRI) and VERDICT MRI. Among these, 44 underwent targeted biopsy and were analysed with rVERDICT using deep neural networks for fast fitting (∼60 times faster than non-linear least squares minimisation approach). A convolutional neural network classifier assessed the rVERDICT parameters in differentiating Gleason grades measured with accuracy, F1-score and Cohen’s kappa. To assess repeatability, five men were imaged twice.

**Results:** There were 37 cancer lesions: 6 Gleason 3+3, 18 Gleason 3+4, and 13 Gleason ≥4+3. The rVERDICT intracellular volume fraction f_ic_ discriminated between Gleason 3+3 and 3+4 (p=0.003); Gleason 3+4 and ≥4+3 (p=0.040); and between 5-class Gleason grades with (accuracy, F1-score,kappa)=(8,7,3) percentage points higher than classic VERDICT, and (12,13,24) percentage points higher than the ADC from mp-MRI. Repeatability of rVERDICT parameters was high (R^2^=0.79–0.98,CV=1%–7%,ICC=92%-98%). T2 values estimated with rVERDICT were not significantly different from those estimated with an independent multi-TE acquisition (p>0.05).

**Conclusion:** rVERDICT allows for accurate, fast and repeatable estimation of diffusion and relaxation properties of PCa and enables discriminating Gleason grade groups.

## Introduction

As for many cancers, definitive prostate cancer (PCa) diagnosis relies on biopsies^1^. This invasive procedure can have serious side effects, such as infection, bleeding and urinary retention, significantly impacting quality of life^2,3^. Recent advances in medical imaging have played a key role in improving PCa detection. For instance, multi-parametric MRI (mp-MRI), consisting of T2-weighted, diffusion-weighted and dynamic contrast-enhanced imaging sequences has been incorporated into the National Institute for Health and Care Excellence (NICE) guidelines for PCa diagnosis^4^. However, whilst mp-MRI has a 90% sensitivity for detection of significant cancer, it’s specificity is moderate at 50%^5^; resulting in 1 in 2 men still needing to undergo an unnecessary biopsy^2^. Significant cancer is generally defined by the presence of Gleason pattern 4 tumour within a biopsy^6-9^. Reliably separating lesions on mp-MRI that contain Gleason pattern 4 disease from those with non-significant cancer (Gleason 3+3) or no cancer remains an unmet clinical need.

To address this, microstructure imaging techniques based on diffusion-weighted MRI (DW-MRI)^10-12^ offer sensitivity and specificity to microstructure changes that correspond to different Gleason grades, well above the simple apparent diffusion coefficient (ADC), conventionally acquired as part of standard mp-MRI protocols^13^. In particular, the Vascular, Extracellular and Restricted Diffusion for Cytometry in Tumours (VERDICT) technique^11,14^ was one of the first showing histological specificity both ex-vivo and in-vivo (clinically and preclinically)^11,14-16^. Preliminary results from the clinical trial INNOVATE^17^ reveal that the VERDICT intracellular volume fraction (f_ic_) can discriminate between Gleason 3+3 and 3+4 lesions (p=0.002, AUC=0.93).

However, VERDICT is currently limited in estimating only diffusion parameters without accounting for the inherent relaxation properties of the tissue^18-26^. This leads to questionable accuracy of microstructural parameters, which likely limits their sensitivity. Relaxometry parameters such as T2 relaxation time have also shown capability to discriminate Gleason grades 3 and 4^18,21,22^. Works exploiting joint relaxation-diffusion analysis^26-36^ have shown that these two types of parameters often contain complementary information.

In this work, we hypothesise that a unifying model capturing both relaxation and diffusion effects can enhance the accuracy of both types of estimates and enable Gleason grade discrimination. We propose a new relaxation-VERDICT (rVERDICT) model to estimate jointly the diffusion and relaxation parameters in prostate while harnessing recent developments in machine learning to reduce the model parameters inference times and enhance the potentiality of the rVERDICT for a 5-class Gleason grade classification.

This work capitalizes on the VERDICT imaging protocol (feasible on clinical scanners) and exploits joint relaxation-diffusion analysis using machine learning, providing two technical innovations:

I. a new approach of modelling VERDICT data which accounts for compartment-specific relaxation times, exploiting all the information available from the multi-TE DW-MRI VERDICT acquisition;
II. a deep learning framework for (a) fast, accurate and precise estimation of the rVERDICTparameters and (b) a 5-score Gleason grade classification.

We demonstrate the new rVERDICT comparing our results with the classic VERDICT and ADC from mp-MRI of the same patients. We show that the new framework enables the on-the-fly microstructure imaging of the prostate and Gleason grade discrimination.

## Methods

This study was performed with local ethics committee approval embedded within the INNOVATE clinical trial^17^. The trial is registered with ClinicalTrials.gov identifier NCT02689271.

### Patient population and study design

72 men (median age=64.8 years; range=49.5–79.6 years) were recruited and provided informed written consent. The inclusion criteria were: (1) suspected PCa or (2) undergoing active surveillance for known PCa. Exclusion criteria included: (1) previous hormonal, radiation therapy or surgical treatment for PCa and (2) biopsy within 6 months prior to the scan. All patients underwent mp-MRI in line with international guidelines^37^ on a 3T scanner (Achieva, Philips Healthcare, Best, Netherlands) supplemented by VERDICT DW-MRI (the clinical DCE part of mp-MRI was performed last after the VERDICT MRI).

After the clinical mp-MRI and VERDICT DW-MRI, 44 participants underwent targeted transperineal template biopsy of their index lesion as clinically indicated. The index lesion was defined as the highest scoring lesion identified on mp-MRI with Likert scores (3-5).

The mp-MRI was used to guide cognitive targeted template biopsy (performed by experienced urologists). Specialist genitourinary pathologists (A.F. and M.R) evaluated histological specimens stained with haematoxylin and eosin from the biopsy cores and assigned each biopsy core a Gleason grade. Figure 1 presents a participation flow diagram.

**Figure 1.**
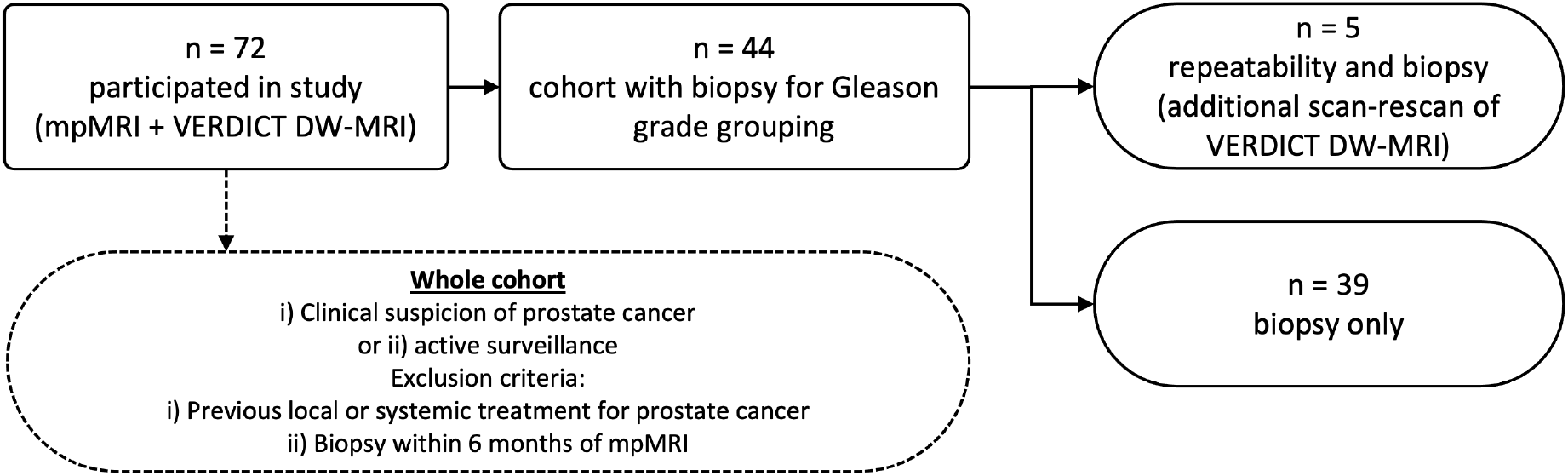
Participation flow diagram. mp-MRI = multiparametric MRI.

### MRI data acquisition

#### DW-MRI acquisition

The VERDICT protocol, adapted from^38^, acquires DW-MRI data using pulsed-gradient spin echo (PGSE) at five combinations (b;δ;Δ;TE;TR) of b-values b (in s/mm^2^), gradient duration δ, separation Δ, echo time TE and repetition time TR (in ms): respectively, (90;3.9;23.8;50;2482); (500;11.4;31.3;65;2482); (1500;23.9;43.8;90;2482); (2000;14.4;34.4;71;3945); (3000;18.9;38.8;80;3349), in three orthogonal directions using a cardiac coil. For each combination, a separate b=0 image was acquired, providing a total of ten different measurements. For b<100 s/mm^2^ the number of averages (NAV)=4 and for b>100 s/mm^2^ NAV=6; voxel size=1.3×1.3×5 mm; matrix size=176×176; average signal-to-noise ratio (SNR)=35; scan duration=12’25”.

#### Repeatability study

Scan-rescan repeatability study of the VERDICT DW-MRI acquisition protocol was performed in five participants (median age=68 years; range=50–79 years). These were randomly chosen among the first 40 participants recruited for the INNOVATE study^17^, thus sharing the same inclusion/exclusion criteria. Participants were imaged twice, taking them out of the scan with less than 5-minute break in between the scans (Figure 1).

#### T2-relaxometry MRI

To additionally assess the performance of rVERDICT in estimating multiple T2 relaxation times in prostate, a multi-TE acquisition was also acquired for an independent estimate of the multiple T2 relaxation times for seven participants (median age=65 years; range=49–79 years), randomly chosen among the participants recruited for the INNOVATE study^17^, thus sharing the same inclusion/exclusion criteria. Details in Supporting Information.

### Image Analysis

#### DW-MRI pre-processing

The DW-MRI pre-processing pipeline included denoising using MP-PCA^39^ with MrTrix3^40^ ‘dwidenoise’; correction for Gibbs ringing^41^ with custom code in MATLAB (The Mathworks Inc., Natick, Massachusetts, USA); correction of motion artefacts and eddy current distortions by mutual-information rigid and affine registration using custom code in MATLAB.

#### T2-relaxometry MRI data pre-processing and analysis

The pre-processing pipeline included only registration of the images at each TE to the image at the first TE, using the same mutual-information rigid registration used for the DW-MRI pre-processing. A two-compartments model was fit to the data to estimate slow and fast T2 values (details in Supporting Information).

#### Classic VERDICT

The VERDICT model^11^ is the sum of three parametric models, each describing the DW-MRI signal in a separate population of water from one of the three compartments: S_ic_ comes from intracellular water (including epithelium), modelled as restricted diffusion in spheres of radius R and intra-sphere diffusivity D_ic_=2 μm^2^/ms (value that minimised fitting error averaged over all PZ voxels and in agreement with recent ultra-short diffusion-time measurements^42^); S_ees_ comes from extracellular-extravascular water adjacent to, but outside cells and blood vessels (including stroma and lumen), modelled as Gaussian isotropic diffusion with effective diffusivity D_ees_=2 μm^2^/ms (value that minimised fitting error averaged over all PZ voxels and in agreement with alternative measurements^31,42^); and S_vasc_ arises from water in blood undergoing microcirculation in the capillary network, modelled as randomly oriented sticks with intra-stick diffusivity D_vasc_=8 μm^2^/ms, which also accounts for any intra-voxel incoherent motion effects. The total MRI signal for the multi-compartment VERDICT model is:

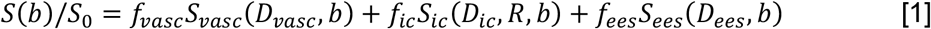

where f_i_ is the proportion of signal from water molecules in population i=vasc;ic;ees, *f*_*vasc*_ +*f*_*ic*_ + *f*_*ees*_ = 1 and *S*_0_ is the b=0 signal intensity. We refer to the original VERDICT work^11,14^ for the specific expressions for S_vasc_, S_ic_ and S_ees_, and the choice and interpretation of the model parameters.

VERDICT has three free model parameters (f_ees_,f_ic_,R) that we estimate by fitting equation [1] to the five DW-MRI measurements at nonzero b values, normalized by their corresponding b=0 measurements. The f_vasc_=1–f_ic_–f_ee_; the cellularity index=f_ic_/R^3^.

The VERDICT model for prostate assumes there are three major tissue compartments that mostly contribute to the measured DW-MRI signal: intra-cellular, intra-vascular and extra-cellular/extra-vascular, and these are non-exchanging (i.e. fully impermeable to water). Several studies^11,14,15^ validated these assumptions under the experimental conditions of the optimized DW-MRI acquisition for VERDICT in prostate^43^. Specifically, VERDICT f_ic_ correlated with only epithelial cells, while the stroma contribution was captured by the extracellular-extravascular compartment^15^. These findings were also supported by an in vivo VERDICT validation study^44^, showing very high correlation (r=0.96, p=0.002) between in vivo VERDICT f_ic_ and epithelial volume fraction from histology.

One of the major limitations of the classic VERDICT formulation is that the signal fractions f_i_ in equation [1] are T2 and T1 relaxation weighted signal fractions. Therefore, their values and interpretation in terms of volume fractions of corresponding tissue compartments can be biased by the unaccounted relaxation properties of the tissue. This occurs when the DW-MRI acquisition includes different TE and TR values for different b values, as in the VERDICT DW-MRI protocol.

#### Relaxation-VERDICT (rVERDICT)

The new rVERDICT model parameterises the T2 relaxation of the intracellular compartment by T2_ic_, and that of vascular and extracellular/extravascular compartments by the same T2_vasc/ees_. It also includes the T1 relaxation contribution from the whole tissue as a single pool. Mathematically, the rVERDICT model is

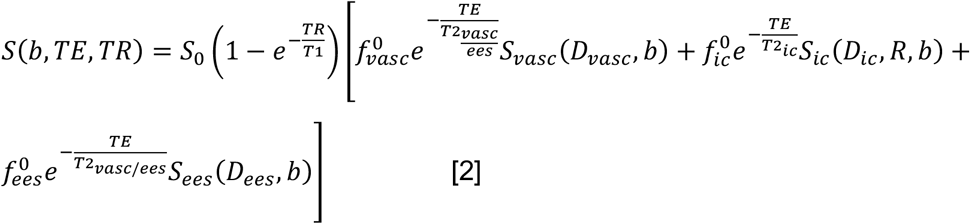

where we adopt the same terminology as VERDICT, but here the volume fractions f^0^_i_, where i=vasc;ic;ees, avoid the bias in the corresponding VERDICT parameters from MR relaxation tissue properties^22^.

rVERDICT has eight free parameters (S_0_,T1,T2_ic_,T2_vasc/ees_,f^0^_ees_,f^0^_ic_,R,D_ees_) that we estimate by fitting equation [2] to the ten DW-MRI measurements: the five nonzero b value measurements and their corresponding five b=0 measurements. Hence, unlike VERDICT, for rVERDICT we exploit the additional TE and TR dependence of the five b=0 measurements to estimate the T2 and T1 relaxation times.

For rVERDICT, the same assumptions as VERDICT for prostate apply, with the additional assumptions about the MR relaxation tissue properties based on currently available experimental evidence^23-25,27,29,30,45-47^. Our choice of a single T1 pool is supported by current literature, showing that it is possible to reliably identify only a single T1 compartment of T1∼[1500–3000] ms^24,25^. Kjaer et al.^23^ also acknowledged that a longer T1 compartment likely exists, but lamented the impossibility to measure it within clinical SNR and time-constraints. For the T2 relaxation, we assume the same T2_vasc/ees_ for the vascular and extracellular-extravascular components. This is supported by previous work which showed that in prostate tissue it is possible to reliably distinguish only two compartments with different T2 values: a slow one, with T2∼[160-1300] ms and a fast one with T2∼[40-100] ms^46^. In our model we assume that the fast T2 compartment is the intracellular space and the slow T2 compartment the vascular (T2 of oxygenated and deoxygenated blood being∼150-250 ms, at 3T and normal hematocrit level∼0.45^47^) and the extracellular-extravascular (typical luminal T2∼[160-1300] ms^46^) space. Stroma is not explicitly modelled.

To investigate the impact of these model assumptions and the stroma contribution (not explicitly modelled), we performed simulations using a four-compartments model accounting for the T2 and ADC values of epithelium, stroma, lumen and vasculature as reported by previous studies at 3T for PCa in the PZ (midgland) in both ex vivo (extrapolated from^30^) and in vivo (extrapolated from^29^). For the vasculature, we used reference values from^38^. Further details in the Supporting Information.

#### Model fitting with deep neural network (DNN)

We obtained quantitative maps from both VERDICT and rVERDICT by fitting respectively equation [1] and [2] to the VERDICT DW-MRI data, using the signal averaged across the three gradient directions. For fast inference, we performed the fitting using a DNN comprised of three fully connected layers^48-50^. We trained the DNN in a supervised fashion using fully synthetic signals generated using equation [1] or [2] with the addition of Rician noise, according to our experimental imaging protocol (Figure 2 and details in Supporting Information).

**Figure 2.**
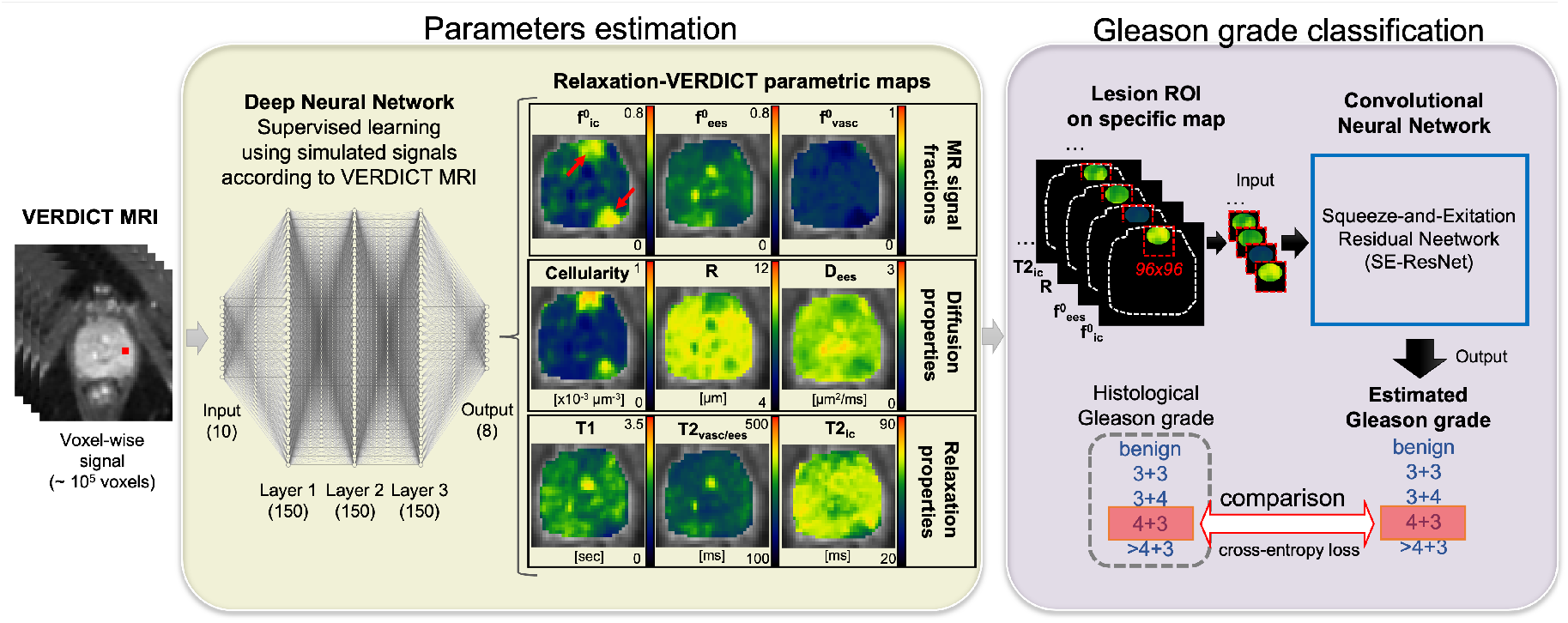
Flowchart of relaxation-VERDICT (rVERDICT) fitting using a fully connected deep neural network and Gleason score classification using a convolutional neural network. We classify the pre-specified lesion ROIs on the rVERDICT parametric maps using a SE-ResNet, whose input is a 96×96 patch with zeros everywhere except within the lesion ROI (dashed red boxes). The model then gives the corresponding Gleason score to the lesion minimizing the cross-entropy loss with respect to the histological grading.

The creation of the training set and training of the DNN (to be done only once) took ∼100 seconds (4 threads on Intel Core i7 processor at 2.4GHz); model parameters prediction for each unmasked DW-MRI dataset (∼5×10^5^ voxels) took ∼35 seconds. For comparison, non-linear least squares minimisation, using the ‘nonlincon’ function in MATLAB, took ∼8000 seconds.

#### DNN model fitting assessment

To assess the accuracy and precision of the DNN estimator for a complex model as rVERDICT we performed numerical simulations with known ground-truth. First, to guarantee generalizability, we simulated signals from model parameters combinations covering the whole parametric space, and not just the subset of realistic prostate tissue combinations (see Supporting Information). The DNN was used to predict the rVERDICT model parameters from these signals and we evaluated accuracy and precision in terms of bias and dispersion of the prediction compared to the known ground-truth. For a realistic combination of the model parameters, mirroring values reported for PCa in PZ and TZ^29,30,38^ (T1=2700 ms;T2_ic_=70 ms;T2_vasc/ees_=530 ms;f^0^_ees_=0.40;f^0^_ic_=0.40;R=8 μm;D_ees_=2 μm^2^/ms), we evaluated the stability of the fit with respect to possible degeneracy and local minima by comparing the distribution of predicted values for each model parameter with the known ground-truth, when the other six parameters were varied taking four values linearly spaced within their biophysically plausible ranges (i.e. 4^6^=4096 different noisy realizations).

As benchmark, we compared the performance of our DNN model with the MATLAB’s ‘nonlincon’ function, using a grid-search algorithm for the initial guess.

#### T2 estimates assessment

To assess the differences between the T2 relaxation times estimated using rVERDICT and the independent multi-TE acquisition, we compared the distribution of T2 values estimated by the two methods for all the voxels within the prostate volume, and corresponding median and 25^th^ and 75^th^ percentiles. Statistically significant differences were assessed by a two-sided Wilcoxon rank sum test.

Given the short maximal TE used in our sequence (90 ms), we also performed numerical simulations to assess the accuracy of the estimation of long T2 values (see Supporting Information).

#### Regions-of-interest definition

Two board-certified experienced radiologists (reporting more than 2,000 prostate MR scans per year) manually placed ROIs on the VERDICT DW-MRI, guided by the standard mp-MRI index lesions and confirmed the ROIs with the biopsy results (further details in the Supporting Information).

### Gleason grade classification

All ROI voxels were assigned to one of the corresponding five different histopathologic categories (i.e. Gleason grade groups) reported by the pathologists: benign, and Gleason grades 3+3, 3+4, 4+3 and ≥4+4.

#### Gleason Grade Differentiation

To assess the ability of rVERDICT to discriminate between Gleason grade groups and compare with VERDICT, analysis of variance with Bonferroni multiple comparisons correction was performed to determine statistically significant differences between four groups: benign, Gleason grades 3+3, 3+4, ≥4+3 (for consistency with previous studies^16^), for all rVERDICT and VERDICT parameters (considering all the ROIs).

#### Gleason grade groups classification with convolutional neural network (CNN)

To assess the potential of combining rVERDICT with deep learning based classification to enhance Gleason grade groups discrimination, we estimated Gleason grade for each ROI with rVERDICT, VERDICT and ADC maps from the mp-MRI (details on the ADC estimation in Supporting Information), using a CNN based on the SE-Res-Net ^51^ trained in a supervised fashion using all the ROIs and the associated five histopathologic categories, i.e. a 5-class classification task (Figure 2).

Specifically, we chose the SE-Res-Net^51^ which gives better performance with our data than DenseNet, another architecture that shown to provide high capacity with multi-layer feature concatenation^52^. More details in^51^ and Figure S1.

For the data imbalance, we used 5-fold cross-validation (with test size 20%) with stratified randomized folds to preserve the percentage of samples for each class. For training, we cropped the images selecting the ROIs’ bounding box to the same size 96×96 pixels. We used 47 ROIs (18 from benign tissue and 29 from cancer lesions) for training and 12 ROIs for testing (4 from benign tissue and 8 from cancer lesions); and applied data augmentation using rigid transformation (affine/rotation). We trained the network for 30 epochs with cross-entropy loss and Adam optimization (learning rate=10^−5^).

### Statistical Analysis

#### Scan-rescan repeatability

We quantified repeatability using the adjusted coefficient of determination R^2^ between each estimated model parameter in the first scan with the estimates from the second scan, considering all the voxels within each ROI (n=179). We used subject-specific ROIs instead of whole prostate statistics to remove potential bias due to deformation and different position of the prostate between the two scans. We used Bland-Altman plots and computed the coefficient of variation (CV) as standard deviation over the mean and the intraclass correlation coefficient (ICC), calculated for two-way mixed effects, single measurement, with absolute agreement.

#### Gleason grade classification performance

We quantified the performance of the 5-class CNN classification by computing the accuracy, precision and sensitivity (recall). From these, we computed the harmonic mean of precision and recall (F1-score) and the Cohen’s kappa coefficient (kappa)^53^.

## Results

There were 37 cancer lesions in the investigated cohort (n=44), and 22 regions that were determined as benign tissue on biopsy. Median prostate-specific antigen (PSA) level was 7.0 ng/mL (range=1.0–71.0 ng/mL), the median time between VERDICT MRI and biopsy was 66.9 days (range=8–167 days). Of the 37 cancer lesions, 6 were Gleason grade 3+3, 18 were 3+4, and 13 were ≥4+3. Table 1 provides a summary of the demographic data.

**Table 1.** Summary of Demographic Data. Note: except where indicated, data are numbers of participants. Numbers in parentheses are ranges. The index lesion was defined as the highest scoring lesion identified on mpMRI with Likert scores (3-5). PSA = prostate specific antigen.

| **Parameter** | **Cohort with biopsy** |
| --- | --- |
| Number of Participants | 44 |
| Median age (y) | 67 (49-79) |
| Median PSA level (ng/ml) | 7.96 (0.83-72.11) |
| Highest Gleason grade of biopsied index lesion |  |
| Benign | 22 |
| 3+3 | 6 |
| 3+4 | 18 |
| ≥4+3 | 13 |
| Median no. of total cores | 23 (6-36) |
| Median no. of sites | 2 (1-13) |
| Median no. of positive cores | 5 (1-15) |
| Median maximum cancer core length (mm) | 8 (1-14) |
| Median maximum cancer core length (%) | 75 (10-100) |
| Median Prostate volume (ml) | 43 (15-108) |

### Repeatability

For the diffusion and T2 relaxation parameters from rVERDICT, R^2^=[0.79-0.98]; CV=[1%-7%]; and ICC=[92%-98%]. The correlation plots and Bland-Altman plots of all the rVERDICT parameters are reported in Figure 3.

**Figure 3.**
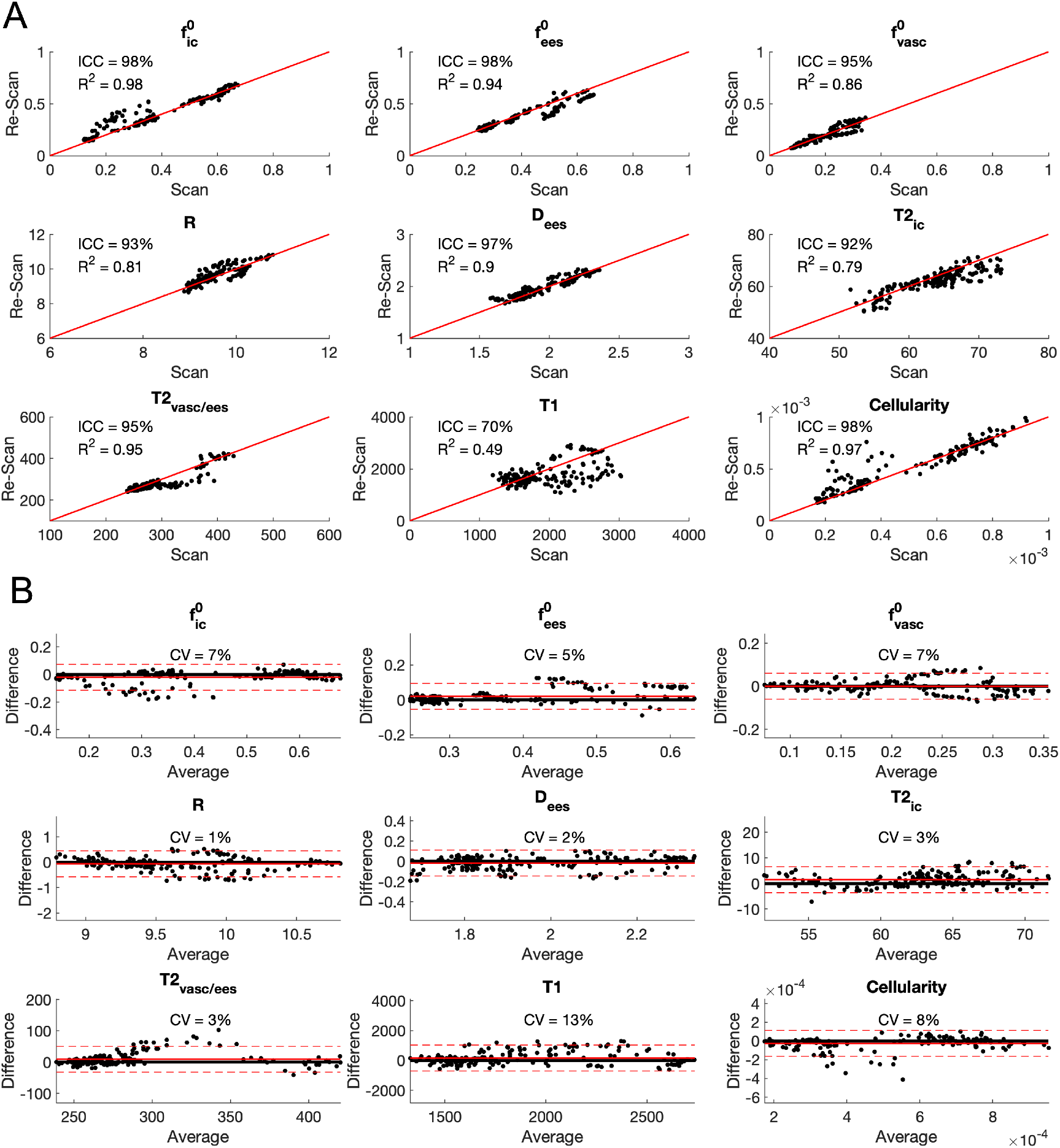
Repeatability of the rVERDICT parameters. A) Correlation plots for all the rVERDICT parameters in the scan/rescan study. The corresponding R^2^ and ICC are reported for each of them, together with the identity line. B) Bland-Altman plots of the scan/rescan estimates for all the rVERDICT parameters. The corresponding CV is reported for each of them, together with the average (straight red line) ± 1.96 standard deviation (dashed red lines) of the difference. The dimensional parameters are in μm (apparent cell radius R); μm^-3^ (Cellularity); μm^2^/ms (extracellular-extravascular apparent diffusivity Dees) and ms (T1, intracellular T2_ic_ and vascular/extracellular-extravascular T2_vasc/ees_).

### DNN model fitting performance

Results from numerical simulations reported in Figure 4 show that the DNN had similar accuracy but higher precision and robustness for all the rVERDICT parameters, compared to conventional non-linear least squares minimisation.

**Figure 4.**
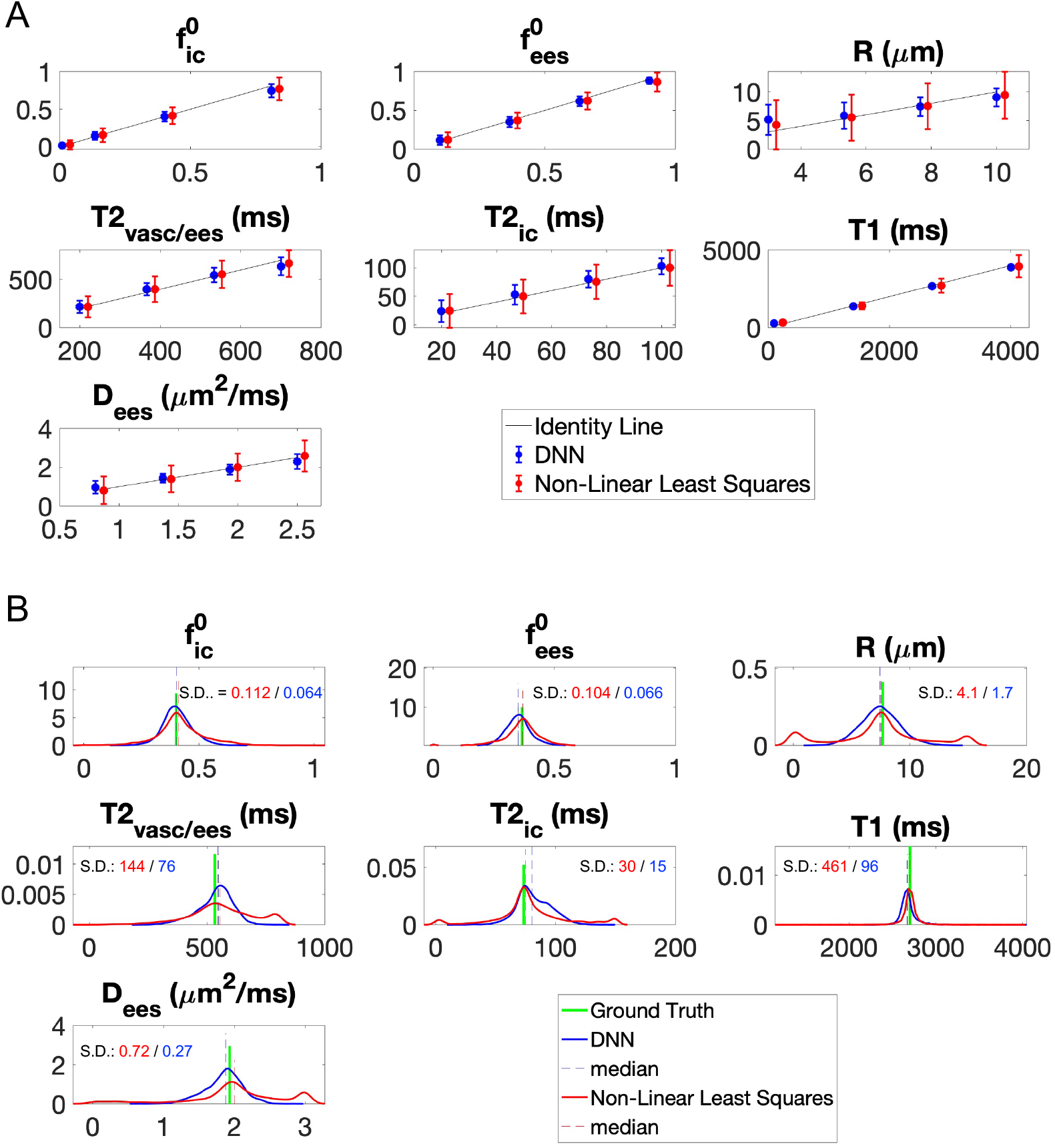
Accuracy and precision of model fitting. A) The mean (data points) and variance (error bars) of the prediction for DNN and conventional non-linear least squares optimization are plotted against the known ground truth from numerical simulations. The identity line is also plotted to aid appreciating the accuracy of the prediction from both methods (higher the accuracy, closer the mean prediction to the identity line). The variance of the prediction (error bars) is a good indicator of the precision of the estimation: smaller the variance, higher the precision. To make the results visually clear, data points for the non-linear least squares were purposely moved slighted to the right. B) The probability density distribution of the estimates of the seven rVERDICT model parameters (S0 was fixed to 1) are plotted for seven ground truth values representative of PCa in TZ and PZ and 4,096 different random realizations of the other parameters, for both DNN and conventional non-linear least squares optimisation. The wider the distribution, the less robust the estimation and the lower the precision due to degeneracy and/or spurious minima. We quantified the width of the distributions through their standard deviations (S.D.) reported in each plot with corresponding matching colours.

### Comparison of T2 estimates from rVERDICT and T2-relaxometry MRI

Comparison of the distributions of T2 values estimated with rVERDICT and independent multi-TE acquisition for the best and worst cases (in terms of comparable median values) in our cohort are reported in Figure 5. We found median values not statistically different (p>0.05) between the two methods and similar interquartile ranges. The estimated compartmental T2 values are also in agreement with current literature, with T2_ic_∼60 ms and T2_vasc/ees_∼250 ms^23,24,27,29,30,46,47^.

**Figure 5.**
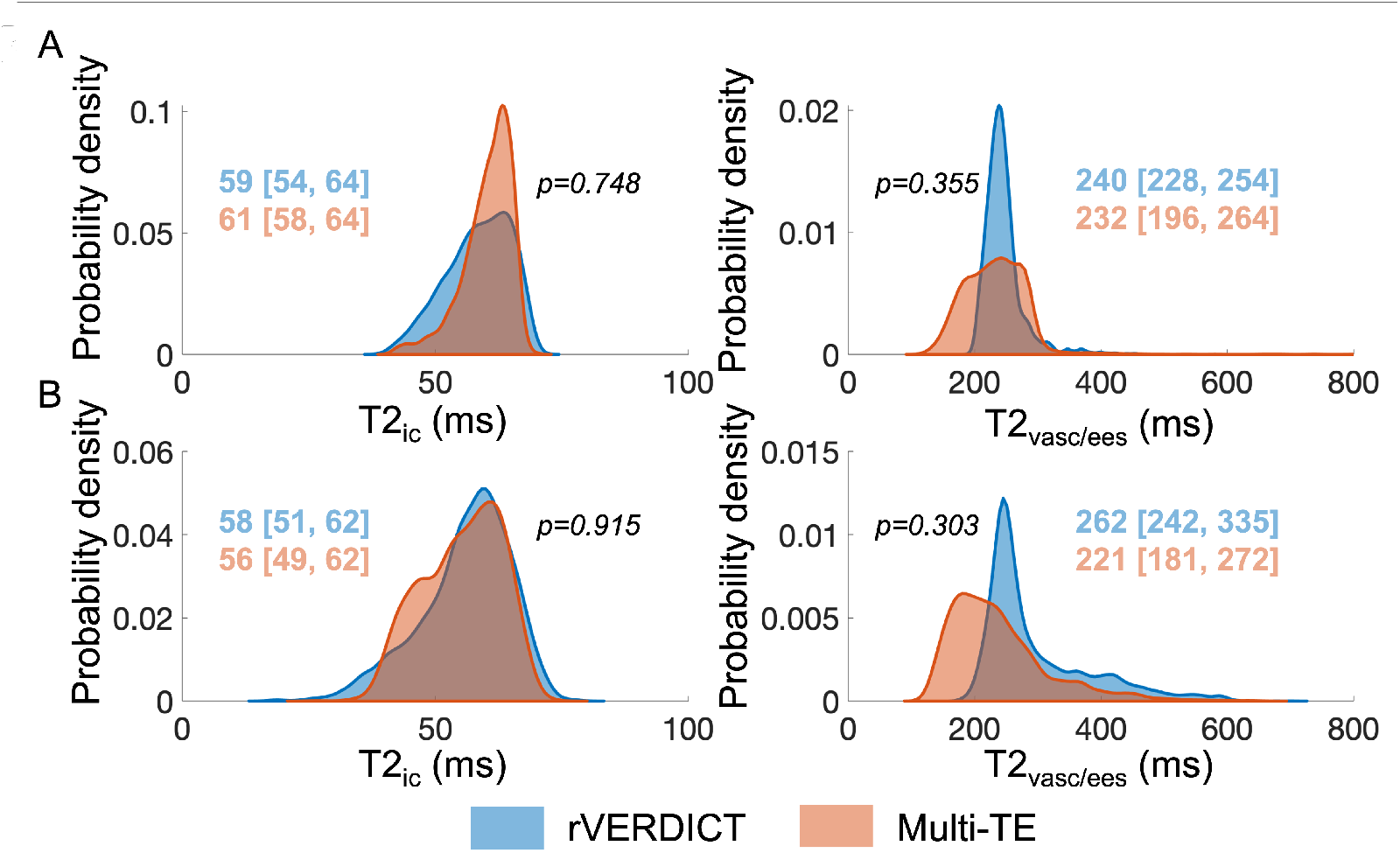
Comparison of T2 estimates from rVERDICT and independent measurements using a multi-TE acquisition. The distributions show the probability density function of the estimated T2 values for all the voxels within the prostate volume for the best (A) and worst (B) cases. Median and [25^th^, 75^th^] percentiles for each distribution are also reported, together with the p values from a two-sided Wilcoxon rank sum test.

### Gleason grades discrimination

Figure 6 reports the box-and-whisker plots of the rVERDICT and VERDICT parameters for the four Gleason grades groups. We found the previously reported ability of VERDICT f_ic_ to distinguish Gleason score 3+3 from 3+4 (p=0.027), but not Gleason 3+4 from ≥4+3 (p>0.05), in agreement with^16^, where the authors additionally showed that ADC does not discriminate between 3+3 and 3+4 (p>0.05), nor 3+4 and ≥4+3 (p>0.05). In contrast, f_ic_ from rVERDICT improved the discrimination of Gleason score 3+3 from 3+4 (p=0.003) and additionally showed promises in discriminating Gleason 3+4 from ≥4+3 (p=0.040). Noteworthy, f_ic_ from VERDICT can distinguish the PCa lesion from benign tissue better than rVERDICT (p=0.017 vs p=0.048).

**Figure 6.**
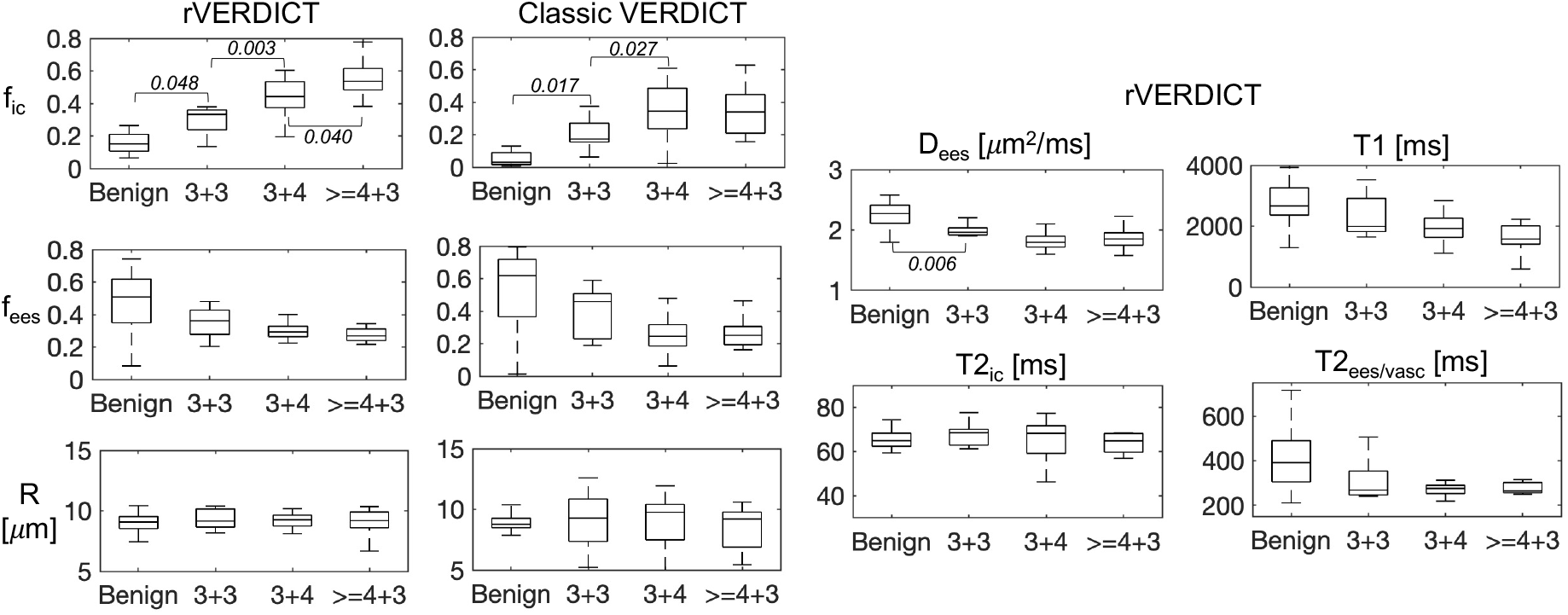
Box-and-whisker plots of the rVERDICT and classic VERDICT parameters as a function of the Gleason grade groups. Only the differences with Bonferroni corrected p<0.05 are considered significant and the corresponding p values reported.

### CNN Gleason grade classification

We obtained the best performances using the 5-class CNN classifier trained with all the rVERDICT parametric maps, achieving accuracy=0.862, F1-score=0.859, kappa=0.815 (the relative contribution of each rVERDICT parameter map is reported in Table S2). For comparison, using the ADC map from mp-MRI we achieved accuracy=0.739, F1-score=0.734, kappa=0.549, while using all the maps from VERDICT we achieved accuracy=0.785, F1-score=0.788, kappa=0.788 (Table 2). The CNN trained and tested only with the intracellular volume fraction f^0^_ic_ map from rVERDICT still outperformed both the CNN trained with f_ic_ from VERDICT and the ADC from mp-MRI (Table 2).

**Table 2.** Overall classification performances. Accuracy, precision, recall, F1-score and kappa for the apparent diffusion coefficient (ADC) maps obtained from multi-parametric MRI (mp-MRI), classic VERDICT and rVERDICT (using only the intracellular volume fraction fic maps or all the parametric maps).

| **Input** | **Accuracy** | **Precision** | **Recall** | **F1-score** | **Kappa** |
| --- | --- | --- | --- | --- | --- |
| ADC mp-MRI | 0.739 | 0.794 | 0.738 | 0.734 | 0.549 |
| $f_{ic}$ classic VERDICT | 0.769 | 0.805 | 0.769 | 0.762 | 0.661 |
| $f_{ic}^0$ rVERDICT | 0.846 | 0.865 | 0.846 | 0.838 | 0.773 |
| All maps classic VERDICT<br>($f_{ic}$, $f_{ees}$, R) | 0.785 | 0.829 | 0.784 | 0.788 | 0.788 |
| All maps rVERDICT<br>($f_{ic}^0$, $f_{ees}^0$, R, $D_{ees}$, $T2_{ic}$,<br>$T2_{vasc/ees}$, T1) | 0.862 | 0.885 | 0.862 | 0.859 | 0.815 |

For per-class Gleason grade classification using all maps from rVERDICT, we found F1-score=0.857, 0.750, 0.889, 0.842, 0.750 for the classes corresponding to benign, Gleason grades 3+3, 3+4, 4+3, >4+3, respectively. All the performances on each Gleason grade classification, using rVERDICT, VERDICT and the ADC from mp-MRI are in Table 3.

**Table 3.**
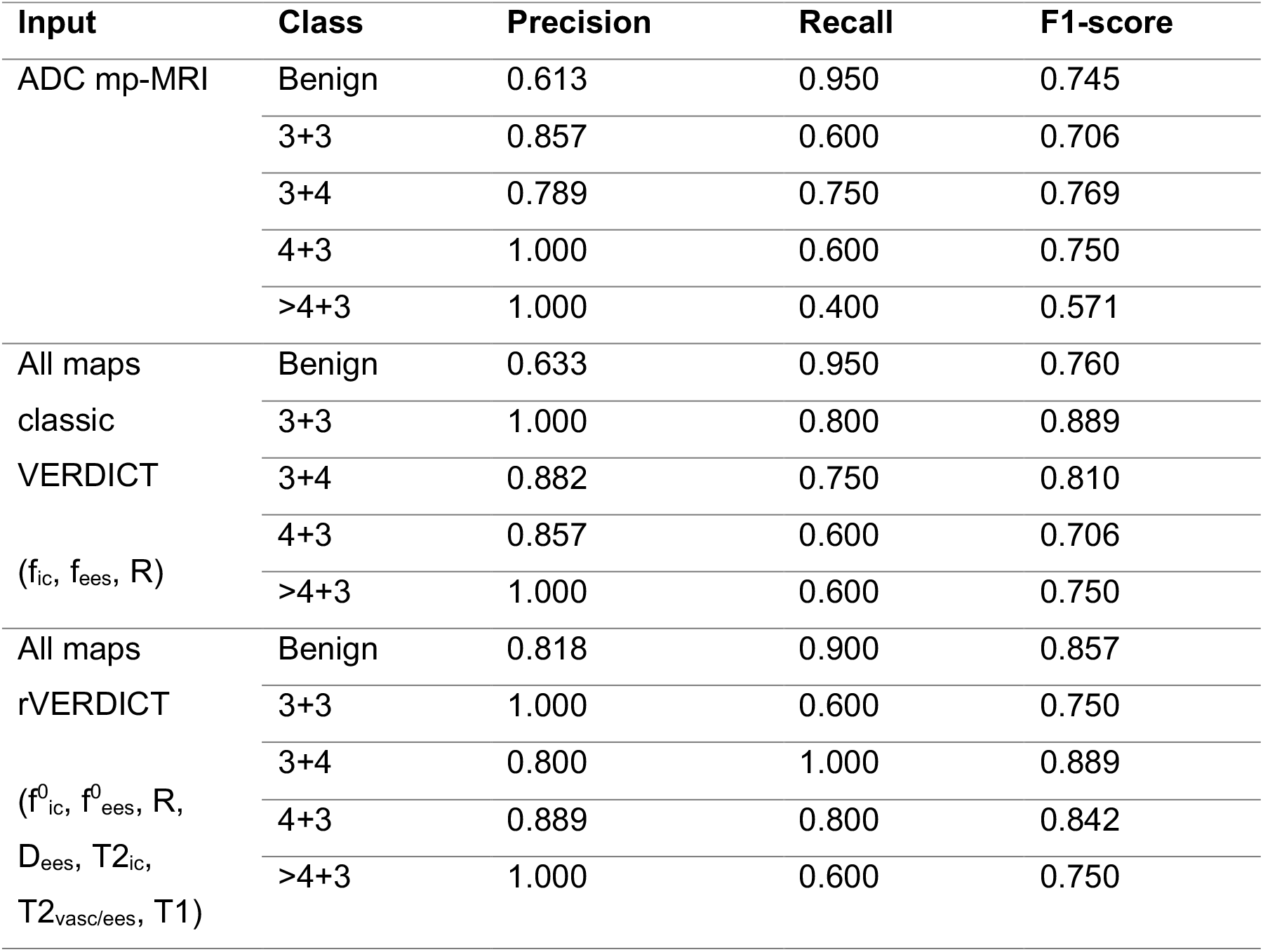
Classification report (precision, recall, and F1-score) for each Gleason grade when using only the apparent diffusion coefficient (ADC) map from multi-parametric MRI (mp-MRI); all the parametric maps from classic VERDICT and all the parametric maps from rVERDICT with 5-class CNN (SE-ResNet).

### rVERDICT parametric maps

Three exemplar cases are shown in Figure 7 to demonstrate lesions with Gleason score 3+3 (Figure 7A, green arrow), 3+4 (Figure 7B, yellow arrow) and 4+3 (Figure 7C, red arrow) on the DWI at b=2000 s/mm^2^, ADC, and rVERDICT f^0^_ic_, D_ees_, T2_vasc/ees_ and T1 maps. A direct comparison of all the VERDICT parametric maps with the corresponding ones from rVERDICT is in Figure S5.

**Figure 7.**
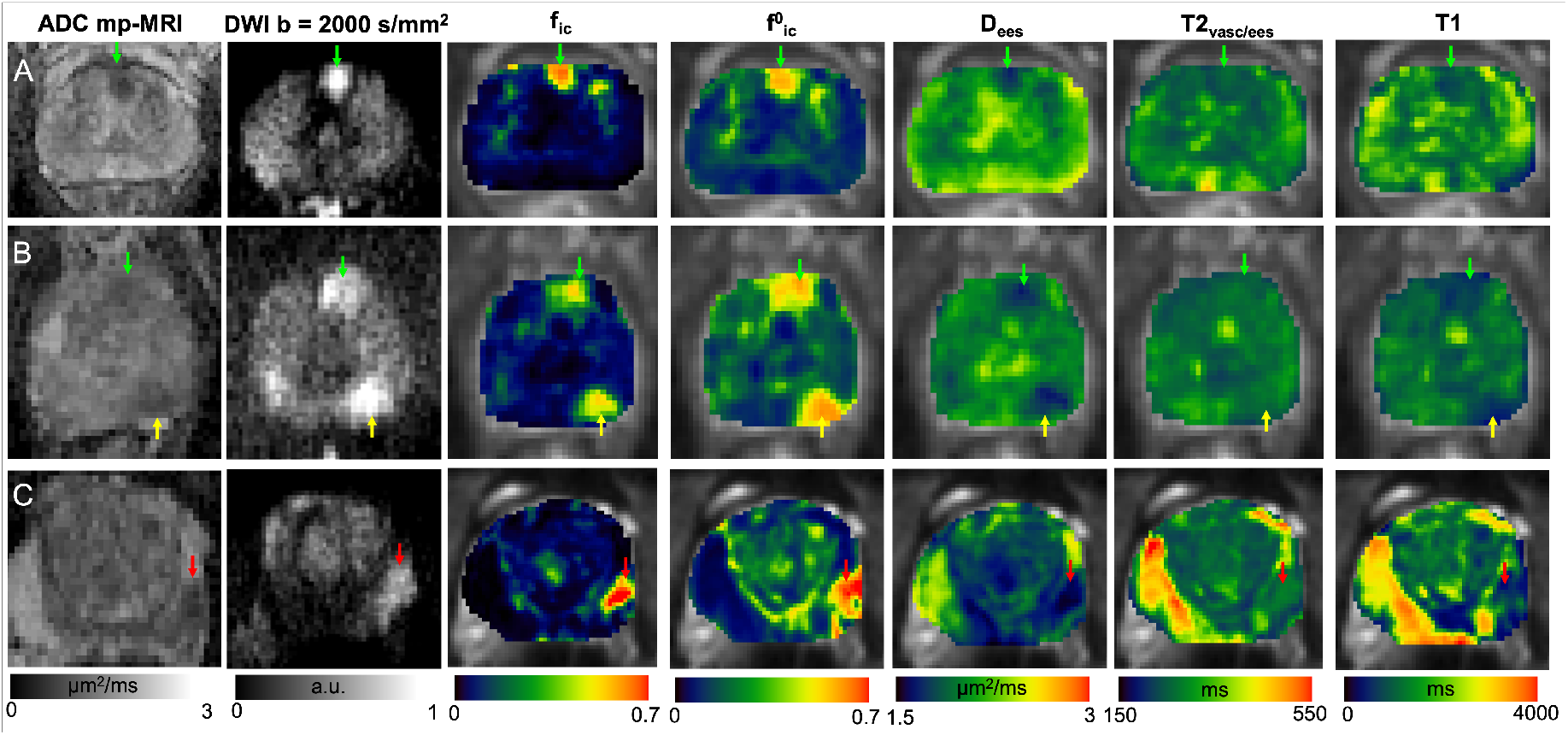
A-C) Apparent diffusion coefficient (ADC) maps from multi-parametric MRI (mp-MRI); diffusion-weighted image (DWI) at b = 2000 s/mm^2^, classic VERDICT intracellular volume fraction f_ic_ and rVERDICT maps (intracellular volume fraction f^0^_ic_; extracellular-extravascular apparent diffusion coefficient D_ees_; vascular/extracellular-extravascular T2 relaxation time T2_vasc/ees_; T1 relaxation time) for three exemplar patients with different PCa: A) age in their 60’s, PSA 4.78 Gleason 3+3 MCCL 7 mm; lesion in the anterior gland; B): age in their 70’s, PSA 5.21 left posterior lesion Gleason 3+4 MCCL 14 mm, left anterior lesion Gleason 3+3; C): age in their 70’s, PSA 8.68 Gleason 4+3, MCCL 10 mm; lesion in the left peripheral zone. Green arrows indicate Gleason grade 3+3, yellow arrows Gleason grade 3+4 and red arrows Gleason grade 4+3.

## Discussion

In this work, we propose a new prostate model called relaxation-VERDICT (rVERDICT) that provides joint estimation of relaxation and diffusion parameters, such as the intracellular T2 relaxation time (T2_ic_) and the intracellular volume fraction (f_ic_). Our hypothesis is that a unifying model capturing both relaxation and diffusion effects would enhance the accuracy of model parameters estimation and consequently the Gleason grade discrimination. As prostate histological components differ between Gleason grades^54^, we expect diffusion parameters, and in particular the f_ic_ from classic VERDICT that correlates with epithelial volume fraction^15,44^, to provide high biologic specificity to Gleason grade. However, classic VERDICT can only achieve discrimination of Gleason 3+3 from 3+4^16^. Gleason discrimination for higher scores like 3+4 from ≥4+3 is also important, as 4+3 cancers are associated with a three-fold increase in lethal PCa compared to 3+4 cancers^6^. Here we hypothesise that rVERDICT can compensate for any relaxation-induced bias that may be reducing the accuracy of classic VERDICT estimates, enabling more robust identification and discrimination of Gleason 4 components.

Our results (Figure 6, Table 2 and 3, and Figure S3 and Table S3) support our hypotheses, showing that the new information obtained from rVERDICT enables the discrimination of Gleason 3+3, 3+4 and ≥4+3 and the classification of five Gleason grade groups, with accuracy and Cohen’s kappa 8 and 3 percentage points higher than VERDICT, and 12 and 24 percentage points higher than the ADC from mp-MRI, respectively. Most importantly, on the previously unattainable classification of Gleason grade 3+4 and ≥4+3, rVERDICT achieved statistical significance (p=0.040 for f^0^_ic_) and the highest precision and sensitivity (both ≥80%) compared to VERDICT and ADC from mp-MRI. The improved performance of rVERDICT over VERDICT is probably due to the compensated relaxation-induced biases on the signal fractions and the extra information on the relaxation times. Our numerical simulations showed that rVERDICT reduces the error of classic VERDICT on (f_ic_,f_ees_,f_vasc_) by respectively (65,93,12) percentage points for the ex vivo and (64,83,20) percentage points for the in vivo case (Figure S3). The analysis of the relative contribution of each rVERDICT parameter to the CNN classification further showed that, in addition to the intracellular volume fraction f^0^_ic_, other rVERDICT parameters such as f^0^_ees_, D_ees_, T1 and T2_vasc/ees_ can offer complementary information to aid the classification of Gleason grade groups (Table S3).

In comparison with previous studies, several machine learning methods have been employed for PCa classification^55-58^ using mp-MRI. A recent survey^59^ reports that most machine learning PCa applications use mp-MRI to solve binary classification (no cancer *versus* significant cancer). For the most challenging five-class Gleason grade classification, the best-performing method in a prostate imaging challenge (Prostate-X2)^55^ achieved kappa=0.270, while subsequent developments using CNNs with mp-MRI achieved higher performances: e.g., U-Net^57^ (kappa=0.446); VGG-Net^56^ (kappa=0.473) and ProstAttention-Net^58^ (kappa=0.511). Our CNN classifier outperformed these results, with rVERDICT yielding a 59% improvement in kappa (kappa=0.815) over these other methods.

The clinical utility of rVERDICT is demonstrated by the repeatability and fitting performance results, alongside the application of rVERDICT to data acquired with a clinical scanner that are also part of an existing clinical trial (registered with ClinicalTrials.gov identifier NCT02689271). The f_ic_ from rVERDICT achieved higher repeatability (R^2^=0.98;CV=7%) compared to VERDICT f_ic_ (R^2^=0.83;CV=27%, from^38^), suggesting that we can achieve greater reproducibility by removing confounds through disentangling relaxation from diffusion parameters. Additionally, the fitting approach based on DNN provides accurate and precise estimates for all the rVERDICT parameters with dramatic reduction of the processing time (∼2 minutes vs ∼2 hours using non-linear least squares minimization, with similar stability, Figure S3), enabling on-the-fly rVERDICT map generation. This is a critical point that further enables clinical translation of the technique, as precision, robustness and computational cost are among the main issues that forbid advanced microstructural imaging from clinical adaptation. Moreover, the DNN proposed in this study is highly generalizable as it can be readily (and quickly) trained using simulated synthetic signals from any arbitrary dMRI acquisition protocol.

A great advantage of rVERDICT is the possibility to obtain simultaneously diffusion and relaxation properties of prostate tissue using only a 12-minute DW-MRI acquisition. We showed that estimates of T2 and T1 relaxation times with rVERDICT match those from independent measurements and literature. The T1 relaxation estimates had lower values within tumour lesions than in benign tissue (mean 1576 vs 2754 ms, respectively, p=0.003), in line with estimates obtained using independent T1 measurements^23-25,45^. The T2 relaxation estimates T2_ic_ within tumours were similar for benign tissue (mean 61 vs 67 ms, respectively, p>0.05) and T2_vasc/ees_ within tumour lesions were lower than in benign tissue (mean 300 vs 383 ms, respectively, p=0.023), in agreement with literature^23,24,27,29,30,46,47^. For seven patients in our cohort, we performed an independent multi-TE acquisition which allowed for a direct comparison of the estimated T2 values using the two methods. Results showed that the rVERDICT T2 values were not statistically different (p>0.05) from the independent T2 measurements (Figure 5). Noteworthy, measurements of T2 relaxation times using multi-echo spin-echo acquisitions may be affected by stimulated and indirect echoes^60^. However, our estimated relaxation times for a two-pool model are in agreement with the literature, including studies that use single-echo spin-echo acquisitions^46^ and MR fingerprinting^45^. Therefore, we believe the bias is negligible in our case. Finally, although multi-TE acquisitions have the advantage of offering MR images with higher resolution and less artefacts compared to DW-MRI (which could partly explain the differences between the corresponding distributions in Figure 5), diffusion-based techniques give us unique insight into microstructure.

To demonstrate the potential of rVERDICT for improving PCa diagnosis, we present three example cases in Figure 7. The new information provided by rVERDICT maps can help improve the ability of identifying and distinguishing Gleason grades of PCa lesions in cases where both the ADC map and the high b-value DWI already show clear contrast (Figure 7A) and most importantly when these conventional measures provide ambiguous information (Figure 7B and 7C). The direct comparison of rVERDICT parametric maps with the classic VERDICT counterparts (Figure S5) showed generally higher f_ic_ and lower f_ees_ estimates, especially in the cancerous areas, in agreement with the simulations in Figure S3 and the results in Figure 6. Although the R maps show differences between the two methods (Figure S5), the estimated cell radius from both rVERDICT and VERDICT ranges between ∼8 and ∼11 μm, in agreement with^15,38,42^.

In comparison to recent diffusion-relaxation techniques proposed for prostate tissue characterization^27,29,30^, rVERDICT has several differences. Firstly, rVERDICT (as VERDICT) explicitly models and quantifies the contribution of vasculature, which is instead neglected in^27,29,30^. While the current literature is still controversial about vascularization and cancer aggressiveness for prostate^61-64^, there is general agreement that increased angiogenesis is an important factor in determining tumour development and prognosis^64^. For cases with aggressive cancer, showing significant vascularisation or neovascularisation, the f^0^_vasc_ map could potentially provide higher discriminative power and/or aid early diagnosis. Also, unlike^29,30^ and similarly to^42^, we explicitly model restriction by considering a compartment of water restricted in the intracellular space, accounting for epithelium. We note that in our model, the signal contribution from stroma is likely captured by the extravascular/extracellular compartment (Figure S2). Previous investigations and histological validations have demonstrated the validity of these assumptions^15,44^, showing good agreement between the VERDICT estimated f_ic_ and f_ees_ with histological measurements of epithelium and stroma plus lumen volume fraction, respectively. Moreover, we note that the results reported in^30^ also suggest that the stroma component does not change significantly in prostate cancer. In rVERDICT (as in VERDICT and^42^) we also model the effective apparent size R of the cellular component that is not modelled in^29,30^ and is only indirectly estimated in^27^.

There are several opportunities for further improvement in the future. The analysis presented here was performed on retrospective data with an acquisition protocol optimised for classic VERDICT probing a limited range of TE and TR values. This mostly compromises the sensitivity to long T2 values (T2_vasc/ees_) and the reproducibility of measured T1 values. However, we have shown that our T2 estimates are still in agreement with independent T2 measurements that cover a wider acquisition parameter space. Also, our simulation results in Figure S4 suggest an error in the estimated values of the long T2 within ±5% of the true value. Future work will explore optimization of the VERDICT MRI acquisition to explicitly account for T1 and long T2 relaxation times, and direct comparison with independent measurements of T1. Additionally, this study analysed only 44 patients for whom the biopsy results were available. This resulted in limited/unbalanced Gleason grades groups, hampering the possibility to examine differences in diagnostic performance (e.g. with comparison of areas under the receiver operating characteristic curve). However, we were still able to draw significant differences and demonstrate the potential of rVERDICT. Further study will include rVERDICT analysis on larger cohorts. From a modelling perspective, rVERDICT (as VERDICT) does not account for the effect of exchange and diffusion time dependence of water diffusivity^27,28^. However, the diffusion time used in this study was between 22-36 ms, a range for which previous studies have shown negligible effects due to permeability and time dependence^27,28,65^. The contribution of stroma is not explicitly modelled by rVERDICT and, as in VERDICT, it is assumed to contribute to the extracellular-extravascular compartment. Given recent experimental evidence that T2 values of stroma are closer to epithelium than lumen^29,30^, we assessed using numerical simulations how assuming a unique average T2 for stroma and lumen could affect the accuracy of estimating f^0^_ic_. We found that this assumption leads on average to underestimate the true epithelial signal fraction by ≤20 percentage points (both ex vivo and in vivo). This bias reduces to ≤10 percentage points when high SNR (≥100) can be achieved (Figures S2 and S3). Future work can explore the possibility including these effects in the model and potentially estimating other tissue properties such as cell membrane permeability and isolate and estimate the stroma contribution. Finally, the DNN used for model parameters estimation does not account for any spatial relationship between voxels. Future work may consider using a CNN based architecture which would naturally account for such spatial relationship and regularize the fitting of rVERDICT to the data, perhaps providing some additional benefits in terms of accuracy and robustness to noise.

## Conclusions

In conclusion, rVERDICT with machine learning allows for accurate, fast and repeatable microstructural estimation of both diffusion and relaxation properties of prostate cancer. This enables classification of Gleason grades, potentially allowing the utilisation of rVERDICT for clinical use and improved diagnosis.

## Data Availability

Data were collected within the INNOVATE clinical trial, registered with ClinicalTrials.gov identifier NCT02689271 and they are not publicly available.

## ACKNOWLEDGMENTS

This work was supported by EP/N021967/1, EP/R006032/1 and by Prostate Cancer UK: Targeted Call 2014: Translational Research St.2, project reference PG14-018-TR2. M.P. is supported by the UKRI Future Leaders Fellowship MR/T020296/2.

## Supporting Information

**Figure S1.**
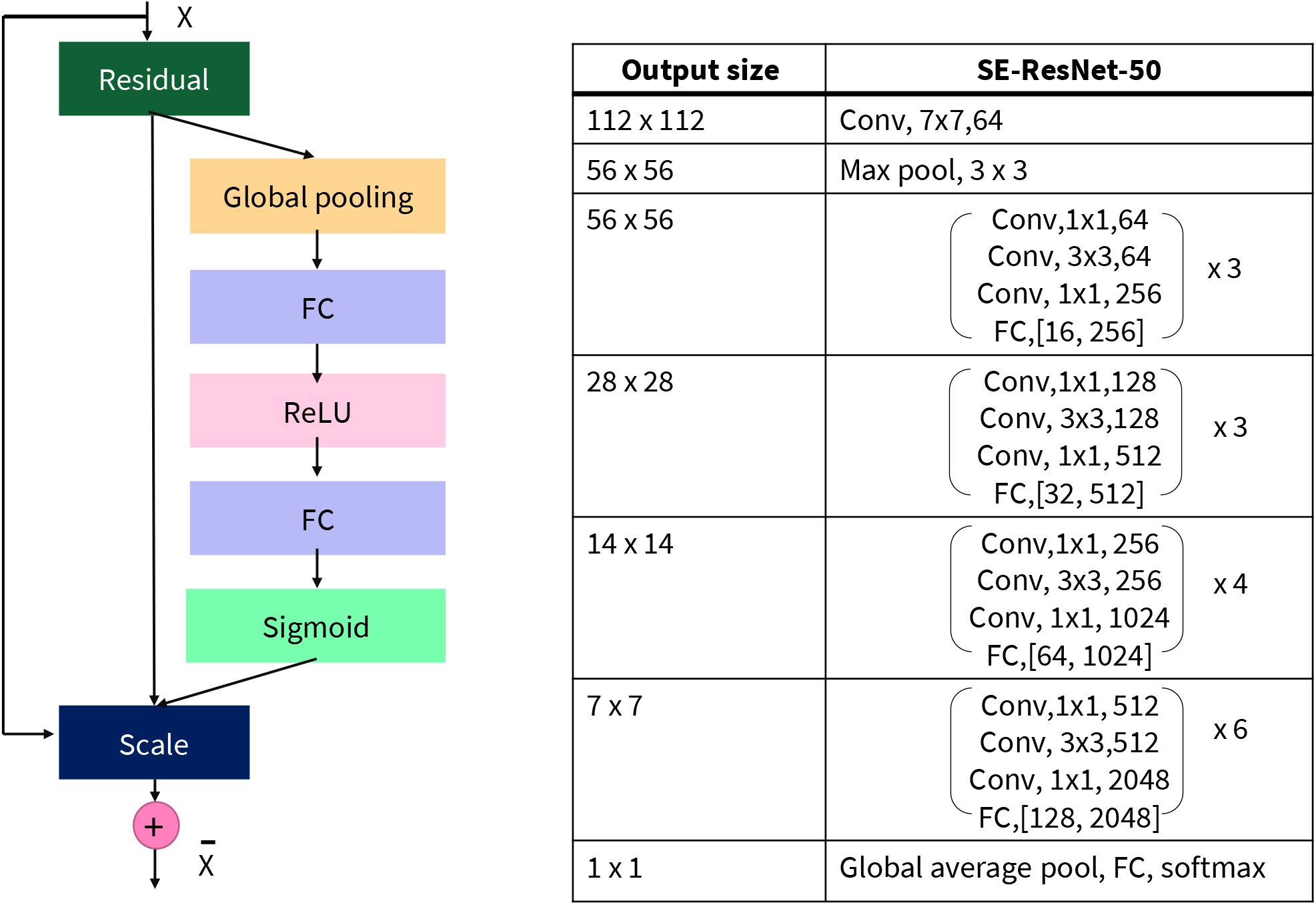
Scheme and details of the specific SE-ResNet architecture used for the 5-class classification of Gleason scores. SE-ResNet combines ResNet with embedded “Squeeze-and-Excitation (SE)” block that adaptively recalibrates feature response. SE blocks intrinsically introduce dynamics conditioned on the input, helping to boost feature discriminability. In SE-ResNet, SE was added before summation with the identity branch.

**Table S1.** Overall classification performances for the chosen CNN architecture, SE-ResNet 50, and the Densenet 264, which showed high capacity with multi-layer feature concatenation^52^. Accuracy, kappa and F1-score for the maps from rVERDICT (using only the intracellular volume fraction f^0^_ic_ maps or all the parametric maps).

| Network | Input | Accuracy | Kappa | F1-score |
| --- | --- | --- | --- | --- |
| Densenet 264 | rVERDICT $f_{ic}^0$ | 0.831 | 0.776 | 0.816 |
|  | All maps rVERDICT | 0.800 | 0.739 | 0.794 |
| SE-ResNet 50 | rVERDICT $f_{ic}^0$ | 0.846 | 0.773 | 0.838 |
|  | All maps rVERDICT | 0.862 | 0.815 | 0.859 |

**Table S2.** The relative contribution of each rVERDICT parameter map for the chosen 5-class CNN classifier, SE-ResNet 50, in terms of Accuracy, kappa and F1-score.

| rVERDICT<br>parameter map | Accuracy | Kappa | F1-score |
| --- | --- | --- | --- |
| $f_{ic}^0$ | 0.846 | 0.773 | 0.838 |
| $f_{ees}^0$ | 0.831 | 0.819 | 0.832 |
| R | 0.800 | 0.668 | 0.802 |
| D<sub>ees</sub> | 0.846 | 0.876 | 0.848 |
| T2<sub>vasc/ees</sub> | 0.800 | 0.700 | 0.797 |
| T2<sub>ic</sub> | 0.815 | 0.683 | 0.817 |
| T1 | 0.876 | 0.837 | 0.874 |

### Conventional ADC measurements

All participants were scanned with a diffusion-weighted echo-planar imaging sequence to estimate the apparent diffusion coefficient (ADC) maps (as part of mp-MRI) with: TR/TE=2753/80 ms; slice thickness 5 mm; no interslice gap; acquisition matrix 168 × 169 mm; b=0, 150, 500, 1000 s/mm2; and six directions per b value. The total imaging time was 5’16”. ADC maps were calculated by using all b values except b=0 to reduce perfusion effects^66^ and were calculated with the Camino Diffusion MRI toolkit^67^.

### Multi-TE T2-relaxometry MRI acquisition

The multi-TE acquisition for independent estimation of T2 relaxation times in prostate consisted of a multi-echo spin-echo sequence with an echo spacing of 31.25 msec and TR=8956 msec. The other imaging parameters were: number of echo times=32; field of view (FOV)=180×180 mm; acquired voxel size=2×2×4 mm; scan duration=5’50”.

### Details on Model fitting with deep neural network (DNN)

In this work, we performed the model fitting using the ‘MLPregressor’ implemented in Python scikit-learn 0.23 (https://scikit-learn.org). The input of our DNN is the signal in each MRI voxel, i.e. a vector whose elements are the DW-MRI signals for each of the ten measurements at the different b, TE and TR combinations. Therefore, each MRI voxel is considered as an independent vector of measurements, and no spatial relationship between voxels is considered, neither during training nor during prediction. The DNN then outputs a vector of eight rVERDICT model parameters (or 3 model parameters in the case of classic VERDICT implementation). The DNN consists of three fully-connected hidden layers with 150 units, each characterised by a linear matrix operation followed by element-wise rectified linear unit function (ReLU), and a final regression layer with the number of output units equal to the number of tissue parameters to be estimated. The DNN is optimised by backpropagating the mean squared error (MSE) between ground truth model parameters and DNN predictions. We performed the optimisation with the adaptive moment estimation (ADAM) method^68^ for 1000 epochs (adaptive learning rate with initial value of 0.001; one update per mini-batch of 100 voxels; early stopping to mitigate overfitting; and momentum = 0.9) on 100,000 synthetic DW-MRI signals (split into 80% for training and 20% for validation). We generate the synthetic DW-MRI signals using equation [2] (or similarly [1]) with different values for the model parameters randomly chosen between biophysical plausible intervals: S_0_ = [0, maximum b=0 intensity x 2], T1 = [10, 4000] ms, T2_ic_ = [1, 150] ms, T2_vasc/ees_ = [150, 800] ms, f_ees_ and f^0^_ees_ = [0.01, 0.99], f_ic_ and f^0^_ic_ = [0.01, 0.99], R = [0.01, 15] μm and D_ees_ = [0.5, 3] μm^2^/ms. Note that we chose a value of 150 ms to separate the short and long T2 components during training, according to the rVERDICT’s assumptions. We also added Rician noise corresponding to SNR = 35 to consider experimental noise effect. For the final parameter computation, we used the DNN at the epoch with minimum validation loss.

### DNN model fitting assessment

To assess the accuracy and precision of the DNN estimator, we generated synthetic DW-MRI signals with equation [2] in the main text and all the possible combinations of the seven model parameters using four values linearly distributed in the intervals: T1 = [10, 4000] ms, T2_ic_ = [1, 150] ms, T2_vasc/ees_ = [150, 800] ms, f^0^_ees_ = [0.01, 0.99], f^0^_ic_ = [0.01, 0.99], R = [0.01, 15] μm and D_ees_ = [0.5, 3] μm^2^/ms. These define a 7-dimensional grid comprised of 4^7^ = 16,384 unique combinations of the seven model parameters and uniformly covering the whole parametric space of rVERDICT model.

### Multi-TE T2-relaxometry MRI analysis

To provide an independent T2 estimation of the relaxation times T2_ic_ and T2_vasc/ees_ assumed in rVERDICT, we fitted the following equation to the multi-TE data:

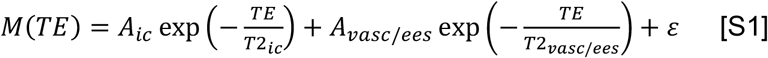

where A_i_ are the relative signal intensities of the compartment i=ic;vasc/ees, T2_i_ are the corresponding T2 values, with T2_ic_<T2_vasc/ees_, and ε is a plateau constant, accounting for non-zero noise floor in magnitude images and found to improve significantly the T2 estimates^46^.

For the fitting of equation [S1] to the T2-relaxometry MRI data, we used the same DNN described in the previous section, trained on 100,000 synthetic signals obtained using equation [S1] with different random values between reasonable intervals: A_i=ic,vasc/ees_ = [0, maximum b=0 intensity x 2], T2_ic_ = [1, 150] ms, T2_vasc/ees_ = [150, 800] ms, and ε = [0.01, 0.20]. We added Rician noise with SNR = 25 to match experimental noise conditions.

### Assessing rVERDICT assumptions for the extracellular-extravascular compartment

In this work we proposed rVERDICT as extension of classic VERDICT to account for compartmental relaxation properties. As such, in rVERDICT we keep the same assumptions as in classic VERDICT. One of these is that the stroma is associated to the extracellular-extravascular compartment. In the original VERDICT work ^11^, this choice was motivated by experimental evidence that the diffusion-weighted MR signal from diffusion restricted in spheres highly correlates with epithelial tissue compartment and it was validated with histology ^15,44^. Since VERDICT is a three-compartment model having one compartment explicitly modelling vasculature, the only other compartment able to account for tissue contributions different from diffusion restricted in spheres (e.g. stroma) is the extracellular-extravascular compartment. Hence, stroma and lumen were assumed to both contribute to the signal of this compartment. Here we performed numerical simulations showing that indeed the overall diffusion-weighted signal decay from stroma is closer to that of lumen, further support this assumption.

We used a complete four-compartment model:

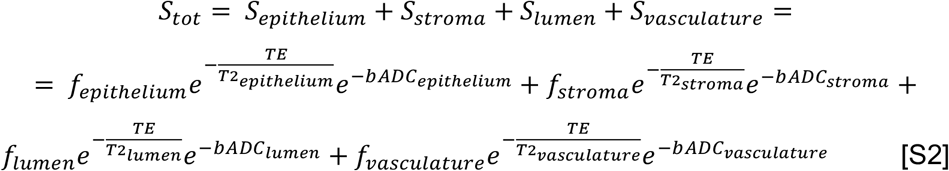

with the TE and b values used in our experiments and the tissue model parameters reported by previous studies at 3T for PCa in the PZ (midgland) in both ex vivo (extrapolated from ^30^) and in vivo (extrapolated from ^29^). For the vasculature component, we used reference values from ^38^. These values are summarized in the table below and the results of the simulated signals are reported in the Figure S2, together with the mean squared error computed between S_epithelium_ and S_stroma_, mse_epithelium-stroma_, and S_lumen_ and S_stroma_, mse_lumen-stroma_.

**Figure S2.**
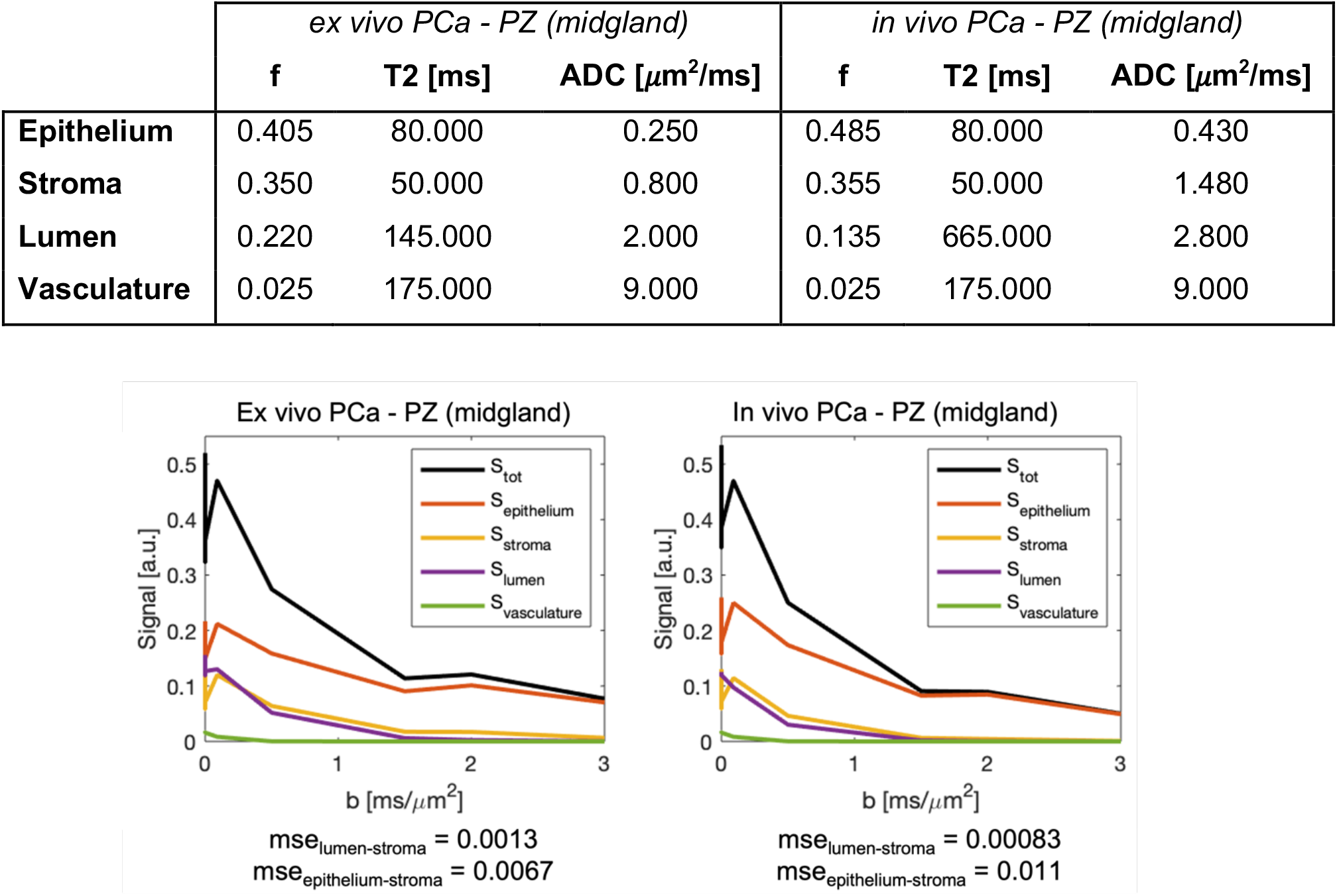
Comparison of the individual signal contributions to the total diffusion-weighted MRI signal (S_tot_) predicted according to the equation [S1] and the protocol used in this study. We focus on S_stroma_ and the assumption of classic VERDICT that its signal signature is more like S_lumen_ (lower mse) than S_epithelium_ (higher mse).

We found that in both the ex vivo and in vivo simulated cases, the signal decay from stroma is closest to that of lumen (i.e., lowest mse), suggesting that, if only three compartments are modelled, then the lowest error can be achieved by coupling the signal from lumen and stroma, as the VERDICT model assumes.

Recent experimental evidence points towards stroma compartment having a T2 value closer to that of the epithelium ^29,30^, we also assessed using these numerical simulations how assuming a unique average T2 for stroma and lumen could affect the accuracy of estimating f^0^_ic_. We have fitted rVERDICT to the S_tot_ we simulated for these two PCa conditions, adding Rician noise with SNR = 35 like in our experiments, and evaluated the average error of our estimates with respect to the ground-truth value of epithelial signal fraction over 1,000 different noisy instances. The results are reported in Supplementary Figure S3. We found that this assumption leads on average to underestimate the true epithelial signal fraction by ∼14 percentage points in the ex vivo case and by ∼20 percentage points in the in vivo case. This bias reduces to ∼7 and ∼10 percentage points, respectively, when high SNR = 100 can be achieved.

However, these results also show that in both cases, rVERDICT (red boxes in Figure S3) reduces the error of classic VERDICT (black boxes in Figure S3) on the estimated signal fractions modelling intracellular (fic), extracellular/extravascular (fees) and vascular (fvasc) compartments by respectively ∼65, ∼93 and ∼12 percentage points for the ex vivo and ∼64, ∼83 and ∼20 percentage points for the in vivo case.

**Figure S3.**
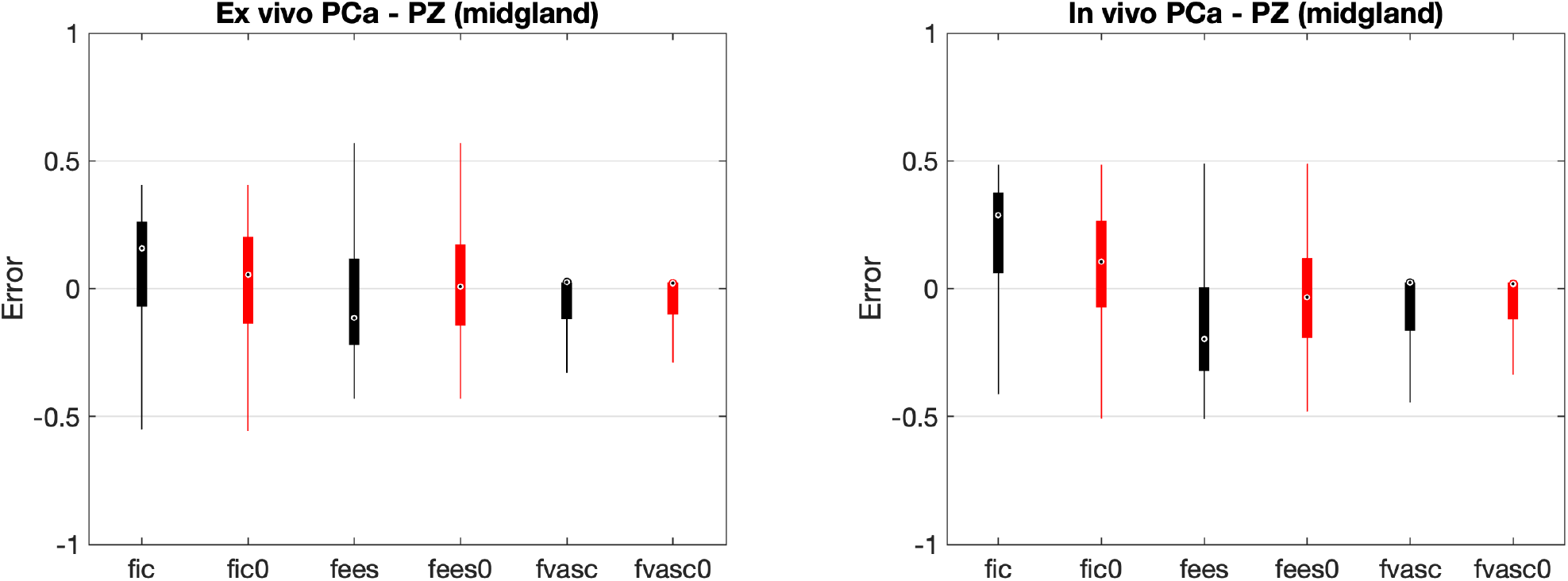
Comparison of the error (i.e. difference with the ground-truth values known by simulation design) in the estimated signal fractions modelling intracellular (ic), extracellular/extravascular (ees) and vascular (vasc) compartments using classic VERDICT (black boxes) and rVERDICT (red boxes), for ground-truth values representative of PCa in the PZ in both ex vivo and in vivo conditions, reported in the table above.

### Accuracy of estimating the long T2 relaxation times

As we have also highlighted in the limitation section in Discussion, the estimates of long T2 components may not be very accurate due to the limited maximal TE used in our sequence (90 ms). To assess how accurate our estimates of long T2 values are, we have performed simulations of a two-compartment system with a short (<=150 ms) and a long (>=150 ms) T2 component using equation [S1] and evaluated the accuracy and precision of the bi-exponential fitting using maximal TE of 90 ms and different SNR by adding corresponding Rician noise. We simulated 30×30×30 = 27,000 different combinations of short and long T2, with different signal fractions of short T2 component, obtained sampling a uniform grid with 30 steps: [signal fraction of short T2, short T2, long T2] = ndgrid(linspace(0,1,30), linspace(1,150,30), linspace(150,800, 30)). The results of these simulations are shown in Figure S4 and they suggest an error in the estimated values of the long T2 within ±5% of the true value for SNR = 35 (case of our experiments).

**Figure S4.**
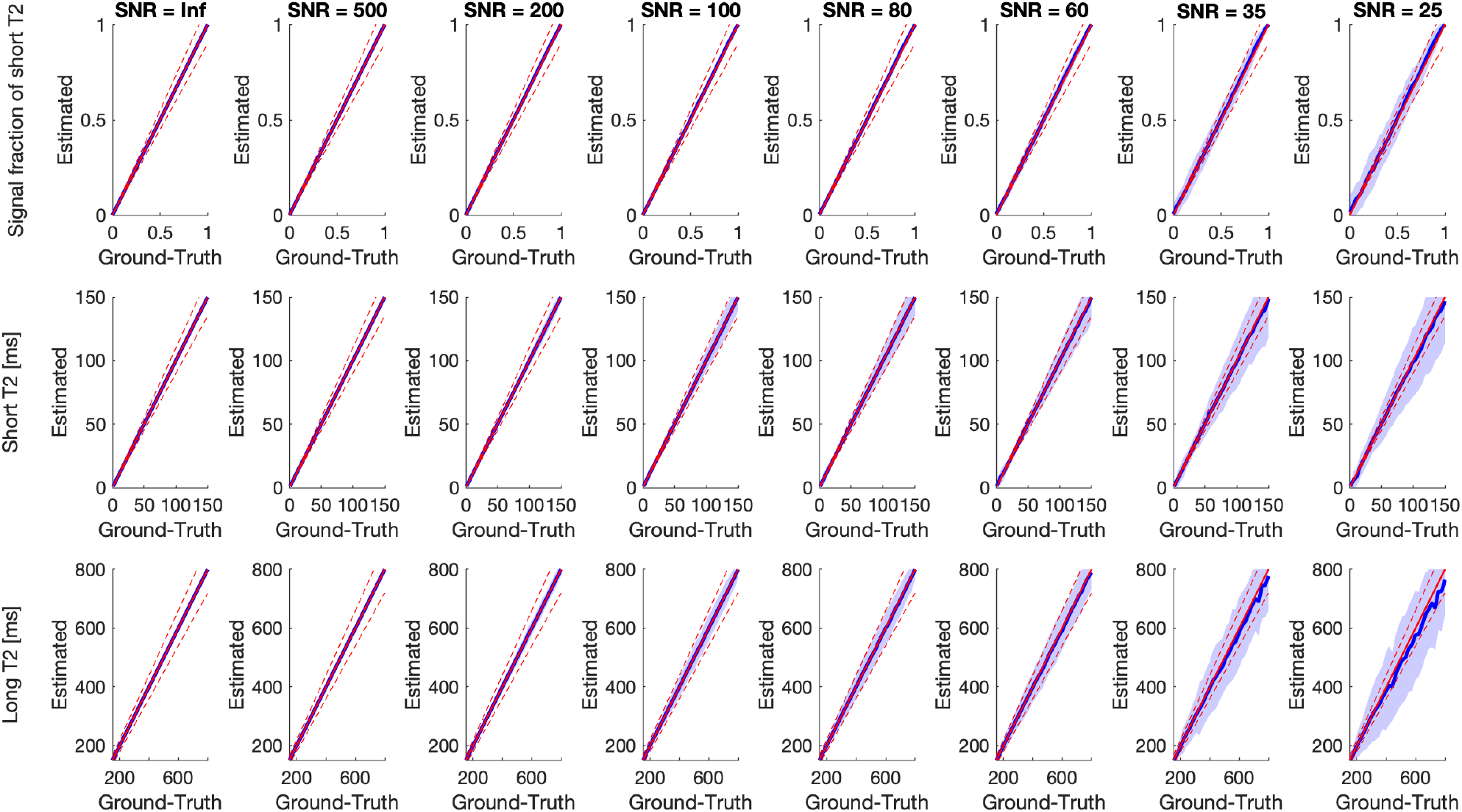
Accuracy and precision of the estimates of the two major T2 components in the prostate tissue using maximal TE of 90 ms, as in our experiments. The red solid lines show the identity lines; the red dashed lines the ±10% interval; the solid blue lines the mean estimates of the three parameters over different noisy instances; the blue shadows the corresponding standard deviation. We found that up to SNR ∼ 60, we have very good accuracy for all the model parameters as well as small standard deviation (i.e. within ±10% of the true value), suggesting high precision. For progressively lower SNR values, the accuracy decreases, with underestimation of the long T2 components for T2 values >∼ 400 ms. However, the error in the estimation of the long T2 component is within ±5% of the true value for SNR = 35 (case of our experiments).

### Additional information on the regions of interest (ROI) definition

The ROIs were informed by the biopsy locations determined by two reads of the participant’s multiparametric MRI by uro-radiologists at our specialist centre. Differences in reports were resolved in a multi-disciplinary meeting. The biopsy targets were indicated on a pictorial report, which was used by a board-certified radiologist to draw ROIs on the lesions on the parameter maps. The radiologist was blinded to histology results. The pictorial reports were used by experienced urologists to carry out MRI-targeted biopsies using ultrasound guidance. MRI targets were matched to real-time ultrasound imaging using cognitive visual registration. This method has been used successfully at our centre for several years and results published in multicenter trials such as the PRECISION prostate trial ^1^. Unexpected biopsy results were also discussed at a multi-disciplinary meeting and repeated if there was a concern for sampling error. We did not include any false positives in this study. Most of the ROIs were 2D. Only in a few cases (less than 6 ROIs) the observable lesions extended to neighbouring slices and in those cases 3D ROIs were considered, but always 2D patches (one for each slice) were fed to the CNN.

**Figure S5.**
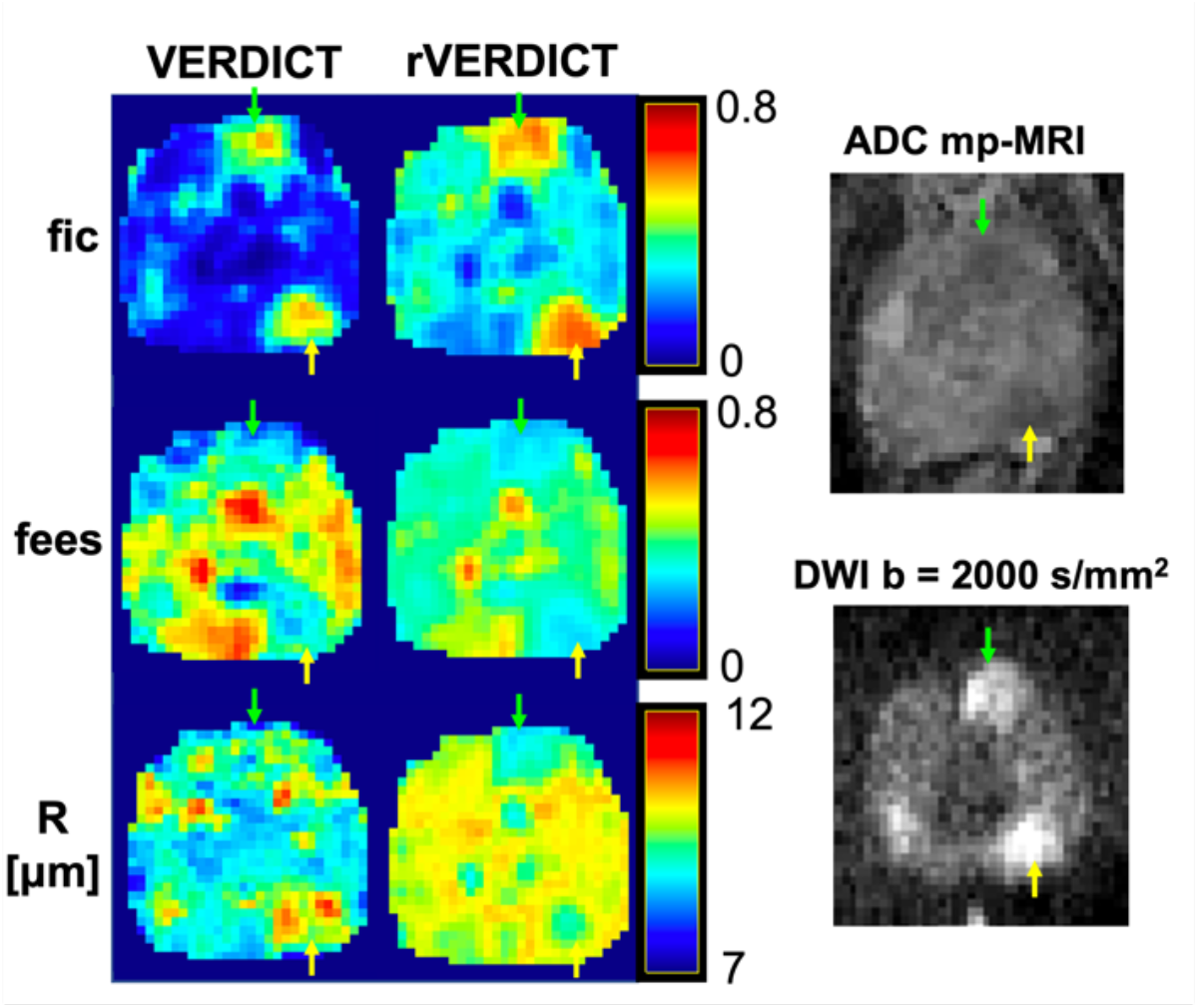
Exemplar comparison of f_ic_, f_ees_, and R maps from classic VERDICT and corresponding ones from rVERDICT for the patient in Figure 6b: age in their 70’s, PSA 5.21 left posterior lesion Gleason 3+4 MCCL 14 mm (yellow arrow), left anterior lesion Gleason 3+3 (green arrow). We observe generally higher f_ic_ estimates, and lower f_ees_ estimates, especially in the cancerous areas, in good agreement with the simulations results reported in Figure S3 and Figure 7 from the main text. Regarding R, the estimates of cell radius from both rVERDICT and VERDICT ranges from ∼8 to ∼11 μm, but their spatial distribution over the whole prostate tissue is different between the two methods.

